# Prevalence of IgG antibodies to SARS-CoV-2 in Wuhan – implications for the ability to produce long-lasting protective antibodies against SARS-CoV-2

**DOI:** 10.1101/2020.06.13.20130252

**Authors:** Tao Liu, Sanyun Wu, Huangheng Tao, Guang Zeng, Fuling Zhou, Fangjian Guo, Xinghuan Wang

**Author notes:** **Correspondence to: Xinghuan Wang, MD**, Department of Urology, Zhongnan Hospital of Wuhan University, No. 169 Donghu Road, Wuchang District, Wuhan, Hubei 430071, China, **and Fangjian Guo, MD, PhD**, Department of Obstetrics & Gynecology, Center for Interdisciplinary Research in Women’s Health, The University of Texas Medical Branch at Galveston, 301 University Blvd, Galveston, TX 77555-0587, and **Fuling Zhou, MD**, Department of Hematology, Zhongnan Hospital of Wuhan University, No. 169 Donghu Road, Wuchang District, Wuhan, Hubei 430071, China. Contributing equally (Tao Liu, Sanyun Wu, Huangheng Tao). Contributing equally (Fuling Zhou, Fangjian Guo, Xinghuan Wang). **Financial Disclosures**. **Author Contributions:** Dr. X. Wang had full access to all the data in the study and takes responsibility for the integrity of the data and the accuracy of the data analysis. Drs T. Liu, S. Wu and H. Tao contributed equally to the study. Drs F. Zhou, F. Guo, and X. Wang contributed equally as senior authors. Concept and design: F. Zhou and X. Wang. Acquisition, analysis, or interpretation of data: T. Liu, S. Wu and H. Tao, Guang Zeng. Drafting of the manuscript: T Liu, S. Wu, F. Guo. Critical revision of the manuscript for important intellectual content: S. Wu, X. Wang. Statistical analysis: T. Liu, H. Tao.

## Abstract

**Background:** It is to be determined whether people infected with SARS-CoV-2 will develop long-term immunity against SARS-CoV-2 and retain long-lasting protective antibodies after the infection is resolved. This study was to explore to explore the outcomes of IgG antibodies to SARS-CoV-2 in four groups of individuals in Wuhan, China.

**Methods:** We included the following four groups of individuals who received both COVID-19 IgM/IgG tests and RT-PCR tests for SARS-CoV-2 from February 29, 2020 to April 29, 2020: 1470 hospitalized patients with COVID-19 from Leishenshan Hospital, Zhongnan Hospital of Wuhan University, and Wuhan No. 7 Hospital, 3832 healthcare providers without COVID-19 diagnosis, 19555 general workers, and 1616 other patients to be admitted to the hospital (N=26473). COVID-19 patients who received IgM/IgG tests <21 days after symptom onset were excluded.

**Results:** IgG prevalence was 89.8% (95% CI 88.2-91.3%) in COVID-19 patients, 4.0% (95% CI 3.4-4.7%) in healthcare providers, 4.6 (95% CI 4.3-4.9 %) in general workers, and 1.0% in other patients (p all <0.001 for comparisons with COVID-19 patients). IgG prevalence increased significantly by age among healthcare workers and general workers. Prevalence of IgM antibodies to SARS-CoV-2 was 31.4% in COVID-19 patients, 1.5% in healthcare providers, 1.3% in general workers, and 0.2% in other patients.

**Conclusions:** Very few healthcare providers had IgG antibodies to SARS-CoV-2, though a significant proportion of them had been infected with the virus. After SARS-CoV-2 infection, people are unlikely to produce long-lasting protective antibodies against this virus.

**Primary Funding Sources:** Part of the study was supported by National Key Research and Development Program of China (2020YFC0845500). The content is solely the responsibility of the authors and does not necessarily represent the official views of the sponsors.

**Role of the Funder/Sponsor:** The sponsors had no role in the design and conduct of the study; collection, management, analysis, and interpretation of the data; and preparation, review, or approval of the manuscript, and decision to submit the manuscript for publication.

**Data and code availability statement:** Data and analyses codes are available from the corresponding authors on request. All request for raw and analyzed data and materials will be reviewed by the corresponding authors to verify whether the request is subject to any intellectual property or confidentiality obligations. Access will be granted after a signed data access agreement is attained.

## Introduction

Currently, coronavirus disease in 2019 (COVID-19) caused by severe acute respiratory syndrome coronavirus 2 (SARS-CoV-2)^1-4^ has become a global pandemic. The virus was freely transmitted among residents in the communities^5-8^ in Wuhan, China from late November 2019 till several days after the lockdown of the city on January 23, 2020. Most of SARS-CoV-2 infections do not require medical attention^9-11^ and only about 5% COVID-19 cases in China need intensive care^12^. As of May 20, 2020, no effective therapies or vaccines for COVID-19 have been reported. Though, numerous therapeutic approaches are under investigation around the world, including existing anti-human immunodeficiency virus (HIV), hepatitis B virus (HBV), hepatitis C virus (HCV) and influenza virus medications^13^, RNA-dependent RNA polymerase inhibitor Remdesivir^14^, antimalarials (chloroquine/hydroxychloroquine), IL-6 blocker Tocilizumab, serum from recovered COVID-19 patients, and inactivated virus, subunit and recombinant vaccines. Among those therapeutic approaches, Remdesivir is most promising so far, although an underpowered trial failed to demonstrate treatment benefits.^15,16^

The presence of SARS-CoV-2 in COVID-19 patients is usually confirmed using real-time reverse-transcriptase polymerase chain reaction (RT-PCR) method.^17-19^ Due to sample collection method, discrepancies in personnel skills training and low virus load in throat swabs after symptom onset^20^, RT-PCR methods have high false-negative test results^19,21,22^. Several days after symptom onset when infected patients start to seek medical attention, virus load in clinical specimens of upper respiratory tract will become relatively low^20,23^. It often takes two or three repeatedly collected specimens to get a positive test result in COVID-19 patients. SARS-CoV-2 enters respiratory epithelial cells via interactions with angiotensin converting enzyme 2 (ACE2)^24^. The spike (S) protein of SARS-CoV-2 mediates binding of the virus with Spike protein’s receptor ACE2 and promotes fusion of viral and host cell membranes and subsequent virus entry into the host cell. In patients infected with SARS-CoV-2, IgM antibodies are detectable around 7 days post infection and IgG antibodies usually take two weeks to develop.^20,24-26^ Recently, several COVID-19 IgM/IgG rapid tests have been developed around the world^27^. It is reported that in some recovered COVID-19 patients who had two negative RT-PCR tests on nasal or throat swabs taken at least 24 hours apart, segments of virus RNA were still detected in other types of clinical samples, especially in fecal swabs^28^. Additionally, some patients had recurrent positive RT-PCR tests on nasal or throat swabs after recovered from COVID-19.^28^ It still unknown whether COVID-19 patients will develop long-term immunity against SARS-CoV-2 and retain long-lasting protective antibodies after the infection is resolved. Large-scale sero-epidemiological studies are also needed to assess infection attack rates and disease incidence in the population and herd immunity. In this study, we reported the experiences in COVID-19 IgM/IgG testing in Wuhan. We assessed prevalence of IgG antibodies against SARS-CoV-2 in hospitalized patients with COVID-19 from Zhongnan Hospital of Wuhan University and Leishenshan Hospital (set up on an emergency basis to admit COVID-19 patients and managed by Zhongnan Hospital of Wuhan University), and Wuhan No. 7 Hospital, healthcare providers without a confirmed COVID-19 diagnosis working in Zhongnan Hospital of Wuhan University, and people from the general population in Wuhan. Considering that Wuhan was the early epicenter of COVID-19 outbreak, most of these healthcare providers would be inevitably exposed to SARS-CoV-2 during the early days of the outbreak (from late November 2019 to January 20, 2020) when person-to-person transmission was not suspected and little personal protection against this virus was employed among medical personnel, a significant proportion of whom would get infected with the virus.

## Results

### Characteristics of participants

Mean age was 58.7 years in 1470 COVID-19 patients, 37.1 years in 3832 healthcare providers, 41.6 years in 19555 general workers and 53.3 years in other patients (**Table 1**). Hospitalized COVID-19 patients were composed of more older people, while healthcare providers and general workers were mostly young adults. Among COVID-19 patients, median time from symptom onset to IgM/IgG test was 41 days (interquartile range 33-50 days). The last RT-PCR tests for SARS-CoV-2 were positive in only eight COVID-19 patients and none of three patients was tested negative for IgG antibodies to SARS-CoV-2.

**Table 1.** Characteristics of hospitalized COVID-19 patients, healthcare providers without confirmed COVID-19, general workers, and other patients in Wuhan (n=26473). COVID-19 patients: hospitalized patients with COVID-19 pneumonia from Leishenshan Hospital, Zhongnan Hospital of Wuhan University, and Wuhan No. 7 Hospital who received both COVID-19 IgM/IgG tests and RT-PCR tests before being discharged from hospital. Healthcare providers: doctors, nurses, and nursing workers without a confirmed COVID-19 diagnosis working in Zhongnan Hospital of Wuhan University who received both COVID-19 IgM/IgG tests and RT-PCR tests before resuming normal clinical services for patients without COVID-19. General workers: general workers in Wuhan who received both COVID-19 IgM/IgG tests and RT-PCR tests before returning to work. Other patients who received both COVID-19 IgM/IgG tests and RT-PCR tests to screen for SARS-CoV-2 infections before being admitted to Zhongnan Hospital of Wuhan University.

|  | Mean (95% Confidence Interval) or n (%) |  |  |  | P value |
| --- | --- | --- | --- | --- | --- |
|  | COVID-19 Patients | Healthcare Providers | General Workers | Other Patients |  |
|  | (n=1470) | (n=3832) | (n=19555) | (n=1616) |  |
| Age (years) | 58.7 (58.0-59.4) | 37.1 (36.7-37.4) | 41.6 (41.4-41.8) | 53.3 (52.4-54.2) | <0.001 |
| Age group (years) |  |  |  |  |  |
| <30 | 30 (2.0) | 1236 (32.3) | 3960 (20.3) | 154 (9.5) | <0.001 |
| 30-39 | 130 (8.8) | 1265 (33.0) | 6068 (31.0) | 166 (10.3) |  |
| 40-49 | 203 (13.8) | 607 (15.8) | 3688 (18.9) | 239 (14.8) |  |
| 50-59 | 356 (24.2) | 600 (15.7) | 4031 (20.6) | 459 (28.4) |  |
| 60-69 | 430 (29.3) | 114 (3.0) | 1315 (6.7) | 357 (22.1) |  |
| ≥70 | 321 (21.8) | 10 (0.3) | 493 (2.5) | 241 (14.9) |  |
| Sex |  |  |  |  |  |
| Female | 745 (50.7) | 2596 (67.7) | 9735 (49.8) | 708 (43.8) | <0.001 |
| Male | 725 (49.3) | 1236 (32.3) | 9820 (50.2) | 908 (56.2) |  |

### Prevalence of IgG and IgM antibodies to SARS-CoV-2

Prevalence of IgG antibodies to SARS-CoV-2 was 89.8% (95% CI 88.2-91.3%) in COVID-19 patients (**Table 2**) compared to 4.0% (95% CI 3.4-4.7%) in healthcare providers, 4.6% (95% CI 4.3-4.9%) in general workers, and 1.0% in other patients (p all <0.001 for comparing to COVID-19 patients). Only the comparison of IgG prevalence between healthcare workers and general workers was not significant (p=0.39). IgG prevalence increased significantly by age among healthcare providers, and was 2.8% in those <30 years old, 9.6% in those 60-69 years old and 10.0% in those ≥70 years old (p<0.001 for trend). IgG prevalence also increased significantly by age among general workers. Prevalence of IgM antibodies to SARS-CoV-2 was 31.4% (95% CI 29.0-33.7%) in COVID-19 patients, 1.5% (95% CI 1.1-1.8%) in healthcare providers, 1.3 (95% CI 1.1-1.5%) in general workers, and 0.2% (95% CI 0-0.4%) in other patients (**Table 3**).

**Table 2.** Prevalence of IgG antibodies to SARS-CoV-2 among hospitalized COVID-19 patients, healthcare providers without confirmed COVID-19, general workers, and other patients in Wuhan (n=26473).

|  | Prevalence % (95% Confidence Interval) |  |  |  |
| --- | --- | --- | --- | --- |
|  | COVID-19 Patients<br>(n=1470) | Healthcare Providers<br>(n=3832) | General Workers<br>(n=19555) | Other patients<br>(n=1616) |
| All | 89.8 (88.2-91.3) | 4.0 (3.4-4.7) | 4.6 (4.3-4.9) | 1.0 (0.5-1.5) |
| Age group (years) |  |  |  |  |
| <30 | 90.0 (78.6-100.0) | 2.8 (1.9-3.8) | 3.5 (2.9-4.1) | 0 |
| 30-39 | 89.2 (83.8-94.6) | 3.8 (2.7-4.8) | 4.7 (4.2-5.3) | 0.6 (0.0-1.8) |
| 40-49 | 92.1 (88.4-95.9) | 4.4 (2.8-6.1) | 4.6 (3.9-5.3) | 1.3 (0.0-2.7) |
| 50-59 | 92.1 (89.3-94.9) | 5.5 (3.7-7.3) | 4.8 (4.1-5.4) | 0.7 (0.0-1.4) |
| 60-69 | 89.1 (86.1-92.0) | 9.6 (4.1-15.2) | 6.8 (5.4-8.1) | 1.1 (0.0-2.2) |
| ≥70 | 86.9 (83.2-90.6) | 10.0 (0.0-32.6) | 5.9 (3.8-8.0) | 2.1 (0.3-3.9) |
| Sex |  |  |  |  |
| Female | 90.3 (88.2-92.5) | 3.7 (3.0-4.4) | 5.0 (4.5-5.4) | 1.0 (0.3-1.7) |
| Male | 89.2 (87.0-91.5) | 4.8 (3.6-6.0) | 4.3 (3.9-4.7) | 1.0 (0.3-1.6) |

**Table 3.**
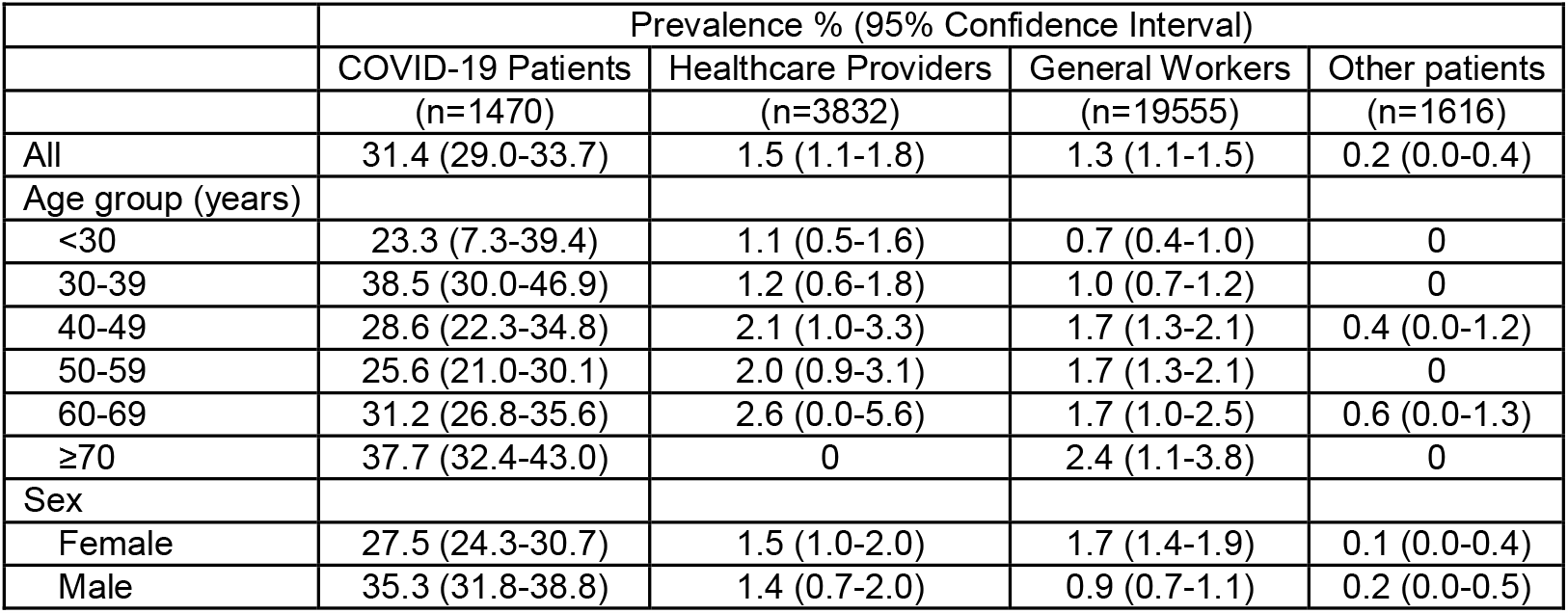
Prevalence of IgM antibodies to SARS-CoV-2 among hospitalized COVID-19 patients, healthcare providers without confirmed COVID-19, general workers, and other patients in Wuhan (n=26473).

### IgM and IgG antibodies to SARS-CoV-2 and mortality in COVID-19 patients

Among COVID-19 patients, mean age was similar between those with IgG antibodies to SARS-CoV-2 and those without (**Table 4**). Presence of IgG antibodies to SARS-CoV-2 was not associated with most demographic characteristics, disease severity, presence of comorbidities, treatment received, and clinical characteristics, except for antibiotics treatment, chloroquine/hydroxychloroquine treatment, and needing intubation. IgG prevalence and IgM prevalence among hospitalized patients with COVID-19 by demographic and clinical characteristics are presented in **Table 5**. Mortality rate was 1.3% (95% CI 0.7-1.9%) in those with IgG antibodies to SARS-CoV-2 and was 3.3% (95% CI 0.4-6.2%) in those without (**Figure 1**). Mortality risk was similar between those with IgG antibodies to SARS-CoV-2 and those without (adjusted hazard ratio 0.45 95% CI 0.16-1.24, P=0.12).

**Table 4.** Demographic and clinical characteristics of hospitalized patients with COVID-19 pneumonia from Leishenshan Hospital, Zhongnan Hospital of Wuhan University, and Wuhan No. 7 Hospital (n=1470).

|  | Mean (95% Confidence Interval) or n (%) |  |  |
| --- | --- | --- | --- |
|  | Positive<br>(n=1320) | Negative<br>(n=150) | P Value |
| Age (years) | 58.5 (57.8-59.2) | 60.8 (58.3-63.2) | 0.061 |
| Age group (years) |  |  |  |
| <30 | 27 (2.0) | 3 (2.0) | 0.26 |
| 30-39 | 116 (8.8) | 14 (9.3) |  |
| 40-49 | 187 (14.2) | 16 (10.7) |  |
| 50-59 | 328 (24.8) | 28 (18.7) |  |
| 60-69 | 383 (29.0) | 47 (31.3) |  |
| ≥70 | 279 (21.1) | 42 (28.0) |  |
| Sex |  |  |  |
| Female | 673 (51.0) | 72 (48.0) | 0.49 |
| Male | 647 (49.0) | 78 (52.0) |  |
| Severity of COVID-19 at admission |  |  |  |
| Moderate | 1004 (76.1) | 108 (72.0) | 0.51 |
| Severe | 263 (19.9) | 34 (22.7) |  |
| Critical | 53 (4.0) | 8 (5.3) |  |
| Comorbidities |  |  |  |
| Any | 739 (56.0) | 87 (58.0) | 0.64 |
| Hypertension | 390 (29.5) | 50 (33.3) | 0.34 |
| Cardiovascular disease (any) | 418 (31.7) | 55 (36.7) | 0.21 |
| Cerebrovascular disease | 40 (3.0) | 7 (4.7) | 0.32 |
| Diabetes | 165 (12.5) | 23 (15.3) | 0.32 |
| Chronic bronchitis/asthma | 23 (1.7) | 2 (1.3) | 0.71 |
| Treatment received |  |  |  |
| Anti-virus | 594 (45.0) | 65 (43.3) | 0.7 |
| Antibiotics | 566 (42.9) | 91 (60.7) | 0.001 |
| Corticosteroids | 148 (11.2) | 18 (12.0) | 0.77 |
| Chloroquine/hydroxychloroquine | 123 (9.3) | 6 (4.0) | 0.029 |
| Vitamin C | 216 (16.4) | 22 (14.7) | 0.59 |
| Immunoglobulin | 12 (0.9) | 2 (1.3) | 0.64 |
| Traditional Chinese Medicine | 1142 (86.5) | 121 (80.7) | 0.051 |
| Clinical characteristics |  |  |  |
| Progression to critical status* | 71 (5.6) | 13 (9.2) | 0.09 |
| ICU admission | 76 (5.8) | 14 (9.3) | 0.084 |
| Need intubation | 28 (2.1) | 11 (7.3) | 0.001 |
| Need Ventilator | 71 (5.4) | 11 (7.3) | 0.32 |
| Death | 17 (1.3) | 5 (3.3) | 0.065 |

ICU: Intensive Care Unit.
\* 61 patients with a critical status at admission were excluded.

**Table 5.** Prevalence of IgM and IgG antibodies to SARS-CoV-2 among hospitalized patients with COVID-19 from Leishenshan Hospital, Zhongnan Hospital of Wuhan University, and Wuhan No. 7 Hospital (n=1470).

|  |  | Prevalence % (95% Confidence Interval) |  |
| --- | --- | --- | --- |
|  | n | IgG | IgM |
| All | 1470 | 89.8 (88.2-91.3) | 31.4 (29.0-33.7) |
| Age group (years) |  |  |  |
| <30 | 30 | 90.0 (78.6-100.0) | 23.3 (7.3-39.4) |
| 30-39 | 130 | 89.2 (83.8-94.6) | 38.5 (30.0-46.9) |
| 40-49 | 203 | 92.1 (88.4-95.9) | 28.6 (22.3-34.8) |
| 50-59 | 356 | 92.1 (89.3-94.9) | 25.6 (21.0-30.1) |
| 60-69 | 430 | 89.1 (86.1-92.0) | 31.2 (26.8-35.6) |
| ≥70 | 321 | 86.9 (83.2-90.6) | 37.7 (32.4-43.0) |
| Sex |  |  |  |
| Female | 745 | 90.3 (88.2-92.5) | 27.5 (24.3-30.7) |
| Male | 725 | 89.2 (87.0-91.5) | 35.3 (31.8-38.8) |
| Severity of COVID-19 at admission |  |  |  |
| Moderate | 1112 | 90.3 (88.5-92.0) | 28.7 (26.0-31.3) |
| Severe | 297 | 88.6 (84.9-92.2) | 37.4 (31.8-42.9) |
| Critical | 61 | 86.9 (78.2-95.6) | 50.8 (37.9-63.7) |
| Comorbidities |  |  |  |
| Any |  |  |  |
| No | 644 | 90.2 (87.9-92.5) | 32.3 (28.7-35.9) |
| Yes | 826 | 89.5 (87.4-91.6) | 30.6 (27.5-33.8) |
| Hypertension |  |  |  |
| No | 1030 | 90.3 (88.5-92.1) | 31.9 (29.1-34.8) |
| Yes | 440 | 88.6 (85.7-91.6) | 30.0 (25.7-34.3) |
| Cardiovascular disease (any) |  |  |  |
| No | 997 | 90.5 (88.6-92.3) | 31.7 (28.8-34.6) |
| Yes | 473 | 88.4 (85.5-91.3) | 30.7 (26.5-34.8) |
| Cerebrovascular disease |  |  |  |
| No | 1423 | 90.0 (88.4-91.5) | 31.3 (28.9-33.7) |
| Yes | 47 | 85.1 (74.5-95.7) | 34.0 (20.0-48.1) |
| Diabetes |  |  |  |
| No | 1282 | 90.1 (88.5-91.7) | 31.1 (28.6-33.7) |
| Yes | 188 | 87.8 (83.0-92.5) | 33.0 (26.2-39.8) |
| Chronic bronchitis/asthma |  |  |  |
| No | 1445 | 89.8 (88.2-91.3) | 31.2 (28.8-33.6) |
| Yes | 25 | 92.0 (80.6-100.0) | 40.0 (19.4-60.6) |
| Treatment received |  |  |  |
| Anti-virus |  |  |  |
| No | 811 | 89.5 (87.4-91.6) | 31.6 (28.4-34.8) |
| Yes | 659 | 90.1 (87.9-92.4) | 31.1 (27.6-34.7) |
| Antibiotics |  |  |  |
| No | 813 | 92.7 (91.0-94.5) | 30.1 (27.0-33.3) |
| Yes | 657 | 86.1 (83.5-88.8) | 32.9 (29.3-36.5) |
| Corticosteroids |  |  |  |
| No | 1304 | 89.9 (88.2-91.5) | 29.1 (26.7-31.6) |
| Yes | 166 | 89.2 (84.4-93.9) | 48.8 (41.1-56.5) |
| Chloroquine/hydroxychloroquine |  |  |  |
| No | 1341 | 89.3 (87.6-90.9) | 30.8 (28.3-33.3) |
| Yes | 129 | 95.3 (91.7-99.0) | 37.2 (28.8-45.7) |
| Vitamin C |  |  |  |
| No | 1232 | 89.6 (87.9-91.3) | 30.4 (27.8-32.9) |
| Yes | 238 | 90.8 (87.0-94.5) | 36.6 (30.4-42.7) |
| Immunoglobulin |  |  |  |
| No | 1456 | 89.8 (88.3-91.4) | 30.8 (28.5-33.2) |
| Yes | 14 | 85.7 (64.7-100.0) | 85.7 (64.7-100.0) |
| Traditional Chinese Medicine |  |  |  |
| No | 207 | 86.0 (81.2-90.8) | 35.3 (28.7-41.8) |
| Yes | 1263 | 90.4 (88.8-92.0) | 30.7 (28.2-33.3) |
| Clinical characteristics |  |  |  |
| Progression to critical status<sup>a</sup> |  |  |  |
| No | 1325 | 90.3 (88.7-91.9) | 30.0 (27.5-32.4) |
| Yes | 84 | 84.5 (76.6-92.4) | 38.1 (27.5-48.7) |
| ICU admission |  |  |  |
| No | 1380 | 90.1 (88.6-91.7) | 30.5 (28.1-32.9) |
| Yes | 90 | 84.4 (76.8-92.1) | 44.4 (34.0-54.9) |
| Need intubation |  |  |  |
| No | 1431 | 90.3 (88.8-91.8) | 30.8 (28.4-33.2) |
| Yes | 39 | 71.8 (57.0-86.6) | 51.3 (34.9-67.7) |
| Need Ventilator |  |  |  |
| No | 1388 | 90.0 (88.4-91.6) | 30.6 (28.2-33.0) |
| Yes | 82 | 86.6 (79.1-94.1) | 43.9 (32.9-54.9) |
| Death |  |  |  |
| No | 1448 | 90.0 (88.4-91.5) | 31.0 (28.6-33.4) |
| Yes | 22 | 77.3 (58.3-96.3) | 54.5 (31.9-77.1) |

ICU: Intensive Care Unit.
<sup>a</sup> 61 patients with a critical status at admission were excluded.

**Figure 1.**
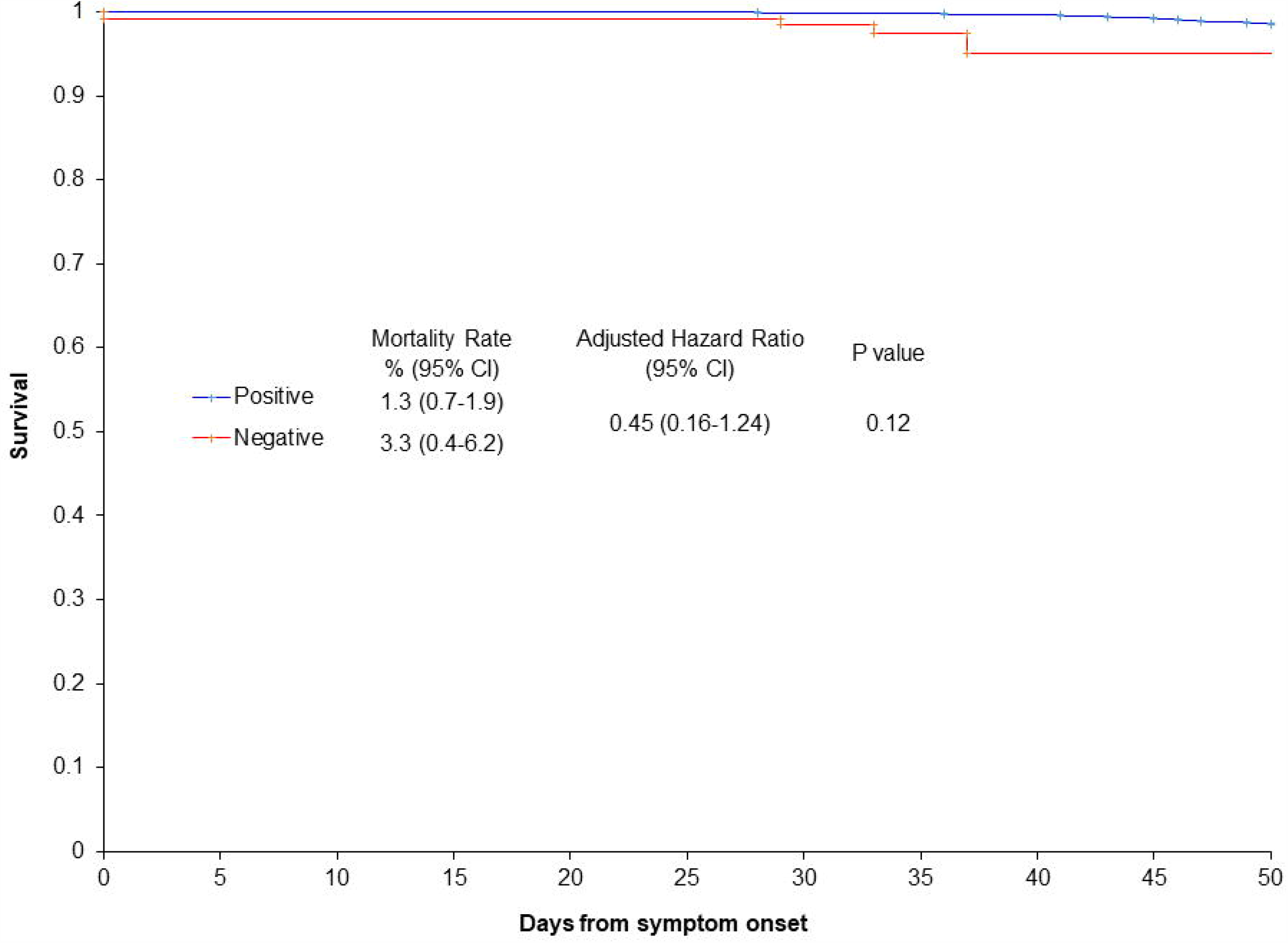
Survival among hospitalized patients with COVID-19 who had IgG antibodies to SARS-CoV-2 and those who did not. Positive: patients who had IgG antibodies to SARS-CoV-2. Negative: patients who did not have IgG antibodies to SARS-CoV-2. Adjust hazard ratio: adjusted for age, sex, and severity of COVID-19 at admission.

## Discussion

We analyzed prevalence of IgG antibodies to SARS-CoV-2 in hospitalized COVID-19 patients, healthcare providers without a confirmed COVID-19 diagnosis, general workers, and other patients to be admitted to hospital. The most intriguing finding of this study is that only 4% of healthcare providers without confirmed COVID-19 diagnosis had IgG antibodies to SARS-CoV-2 in their blood. Most of the healthcare providers were exposed to SARS-CoV-2 during the first few months of the outbreak when use of personal protection equipment was sparse as person-to-person transmission was not suspected. COVID-19 IgM/IgG tests in the US and around the world as reported in the news constantly showed that the true infection rate would be 10 to 80 times higher than that had been confirmed by RT-PCR tests for SARS-CoV-2. Seroprevalence of antibodies to SARS-CoV-2 in 1021 people before resuming work from April 3 to 15, 2020 in Wuhan was reported to be ∼10%,^29^ about 20 times higher than the infection attack rate calculated from the confirmed COVID-19 cases. In New York City, a 21.2% positive rate was reported with young and middle-aged people having the highest positive rate. The proportion of people infected with SARS-CoV-2 who have no symptom or only mild symptoms that do not need medical attention or hospitalization may account for the majority of SARS-CoV-2 infections. Currently, no effective therapeutics are available for treating COVID-19 patients. Detecting patients with SARA-CoV-2 infection who are not in urgent need of hospitalization will provide no benefit to these patients, though it may have important public health applications in tracing their close contacts and preventing these people from spreading the infection to others. In Zhongnan Hospital of Wuhan University, 2.88% (118/4099) healthcare workers were diagnosed with COVID-19 before March 16, 2020. With a moderate estimation, the true infection rate would be ten times that had been confirmed, i.e., >25% of those healthcare providers without diagnosed COVID-19 had been infected. However, only 4% of those infected healthcare workers without confirmed COVID-19 still had IgG antibodies to SARS-CoV-2. They just got infected with SARS-CoV-2 and cleared the virus by their own immune systems. No long-lasting protective antibodies against SARS-CoV-2 were produced in these healthcare providers. Our observed high prevalence of IgG antibodies to SARS-CoV-2 in older groups (60-69 years old and ≥70 years old) among health care workers and general worker in Wuhan also raised the concern that IgG antibodies to SARS-CoV-2 would be lost some time after the infection was cleared, as young or middle-aged people usually took more social responsibilities and had higher chances to get infected during lockdown of the city. We also found that >10% of confirmed COVID-19 cases had no detectable serum levels of IgG antibodies to SARS-CoV-2 after 21 days post symptom onset. They were unlikely to not produce IgG antibodies to SARS-CoV-2 after being infected with the virus^20,24-26,30^. Long et al reported that after 17-19 days post symptom onset, IgG was positive in all patients with COVID-19.^30^ Lack of blood samples >17 days post symptom onset may be responsible for negative IgG tests in some patients reported in previous literatures.^25,26^ Therefore, >10% patients in our study who had no IgG antibodies after 21 days post symptom onset most likely lost those IgG antibodies after the infection was resolved.

After infection with SARS-CoV-1, patients start to produce SARS-specific IgG antibody in the second week, which persists for a long time^31,32^. Even after 210 days after symptom onset, neutralizing viral antibodies (anti-viral IgG) are still detectable in recovered SARS patients^33^. It is believed that the Spike protein and nucleocapsid protein play a central role in the antibody production^31,32,34^. Research has focused on developing vaccines and therapeutics targeting the Spike protein^35,36^ and the nucleocapsid protein^37^. However, our findings indicate that people are unlikely to develop long-lasting neutralizing antibodies to SARS-CoV-2 after SARS-CoV-2 infection. Infections with some viruses, such as HIV, do not illicit robust protective immunity^38^, while common cold coronavirus only generate partial protective immunity^39^. Cross-reactive antibodies in convalescent SARS patients’ sera can neutralize other human betacoronaviruses^40^, as those viruses share a significant B-cell epitope overlapping the heptad repeat-2 region of the Spike protein. SARS-CoV-2 also belongs to betacoronaviruses. However, no individuals or populations had shown explicit immunity to SARS-CoV-2 and every human being in the world was susceptible to this virus before it first jumped from wild animals to humans. It might be due to the dramatic differences between SARS-CoV-1 and SARS-CoV-2 in the receptor-binding region of Spike protein and the key amino acid residues involved in the interaction with human ACE2.^24^ Neutralizing antibodies to SARS-CoV-1 may not produce reliable immunoprotection against SARS-CoV-2. However, why long-lasting protective antibodies are not produced after SARS-CoV-2 infection is still to be studied.

Are IgG antibodies to SARS-CoV-2 parts of short-term immunity against this virus? It is reported that after infected with SARS-CoV-2, rhesus macaques produced antibodies that exhibited neutralizing activity against SARS-CoV-2 in vitro, and re-challenge with the same viral dose weeks post initial infection was not successful.^41,42^ This indicates that short-term neutralizing antibodies may be produced in this animal model. However, infected monkeys have a faster viral clearance mechanism than humans and reinfection protection for humans even in a short time period after the initial infection might not necessarily happen. We found mortality risks were similar between hospitalized COVID-19 patients with IgG antibodies to SARS-CoV-2 and those without, which indicates that absence of IgG antibodies may not affect clinical end outcome and IgG antibodies may even not be part of short-term protective immune response against SARS-CoV-2. Whether therapeutic antibodies targeting the spike protein are effective is also a question. A recent study characterized the viral spike protein receptor-binding domain-specific monoclonal antibodies derived from single B cells of patients with COVID-19.^43^ In that study, the three most server cases (one dead) had much higher plasma binding activities to SARS-CoV-2 spike protein, spike protein receptor-binding domain, and nucleocapsid protein than the other five cases with mild symptoms, which casts some doubts on the relationship between antibody response and disease progression and the utilization of neutralizing antibodies as prophylactic and therapeutic SARS-CoV-2 interventions. Antibody-dependent enhancement of viral entry via the binding of those neutralizing antibodies and the spike protein is also a big concern to be closely monitored^44-47^.

Our findings have important implications for herd immunity, antibody-based therapeutics, public health strategies, and vaccine development. First, as infected people do not develop long-lasting protecting antibodies against SARS-CoV-2, the idea of immune certificate for recovered COVID-19 patients is invalid. This finding also raises concerns for reinfection, chronic infection, and validity of the herd immunity theory for SARS-CoV-2. Second, COVID-19 IgG antibodies as tested by the kits may simply serve as a sign of the infection status and might not be protective neutralizing antibodies. The utilization of convalescent serum from recovered COVID-19 patients in clinical settings^48^ would be questionable. Clinical trials on the efficacy of convalescent serum from recovered COVID-19 patients in treating COVID-19 cases were conducted in Wuhan. Their results will provide further evidence to help elucidate the role of COVID-19 IgG antibodies. Third, as serum COVID-19 IgM/IgG level can become undetectable after recovery in patients with SARS-CoV-2 infections, COVID-19 IgM/IgG test will not be a reliable tool for the surveillance of past infections of SARS-CoV-2 in areas where the epidemic is over. Finally, in our study, most of the healthcare providers without a confirmed COVID-19 diagnosis have been exposed to the wild-type SARS-CoV-2 virus in a highly contagious environment. However, none of them developed long-lasting protective antibodies against SARS-CoV-2. These findings raise the concern if the inactivated virus, subunit, and recombinant vaccines currently under development will be able to induce effective immune protections against SARS-CoV-2.

The main strength of this study is that we analyzed data on COVID-19 IgM/IgG tests on a large cohort of individuals from two hospitals in Wuhan, the epicenter of COVID-19 outbreak in China. Antibody tests on healthcare providers provided valuable information on outcomes of SARS-CoV-2 infections. Limitations of this study include that we do not have long-term follow-up data on recovered COVID-19 patients. Whether or not those patients will lose COVID-19 IgG antibodies in the next few months is still to be studied. Additionally, COVID-19 IgM/IgG test does not have perfect sensitivity and specificity for detecting IgG antibodies to SARS-CoV-2 in the serum. Nevertheless, we observed that very few healthcare providers without a confirmed COVID-19 diagnosis had a positive test result. This observed phenomenon strongly suggests that long-term protective antibodies are unlikely produced after SARS-CoV-2 infection.

In conclusion, very few healthcare providers without confirmed COVID-19 diagnosis in Wuhan have IgG antibodies to SARS-CoV-2, though a substantial portion of them had been infected with the virus. More than 10% of COVID-19 patients did not have those antibodies after 21 days post symptom onset. After SARS-CoV-2 infection, people are unlikely to produce long-lasting protective antibodies against this virus.

## Online Methods

### Study Design and Participants

The study was approved by the institutional ethics board at Zhongnan Hospital of Wuhan University. Requirement for written informed consent was waived by the institutional ethics board for emerging infectious diseases. We included the following four groups of individuals who received both COVID-19 IgM/IgG tests and RT-PCR tests for SARS-CoV-2 from February 29, 2020 to April 29, 2020: hospitalized patients with COVID-19 from Leishenshan Hospital, Zhongnan Hospital of Wuhan University, and Wuhan No. 7 Hospital who received these tests before being discharged from hospital, healthcare providers (doctors, nurses, and nursing workers) without a confirmed COVID-19 diagnosis working in Zhongnan Hospital of Wuhan University who received these tests before resuming normal clinical services for patients without COVID-19, general workers in Wuhan before returning to work, and other patients who received these screening tests before being admitted to Zhongnan Hospital of Wuhan University. There were 1603 patients with COVID-19 who received both COVID-19 IgM/IgG tests and RT-PCR tests for SARS-CoV-2 from February 29 to April 5, 2020 (the last test result was used for analyses). Eighty-four patients were transferred from Zhongnan Hospital of Wuhan University to Leishenshan Hospital and were only counted once each. We excluded 133 patients with COVID-19 whose IgM/IgG tests were less than 21 days after symptom onset to allow enough time for IgG antibodies against SARS-CoV-2 to develop. There were 4099 healthcare providers working in Zhongnan Hospital of Wuhan University, of whom 118 were diagnosed of COVID-19 before March 16, 2020 and 3835 healthcare providers without diagnosed COVID-19 received both tests before resuming normal clinical services. Three healthcare providers who were tested positive for SARS-CoV-2 by RT-PCR tests in their throat swabs were also excluded from the analyses. Before returning to work, 19570 general workers in Wuhan without a diagnosis of COVID-19 who received both tests at Zhongnan Hospital of Wuhan University, of whom 15 were tested positive for SARS-CoV-2 by RT-PCR tests in their throat swabs and were excluded from the analyses. Before admitted to Zhongnan Hospital of Wuhan University for other conditions, 1628 patients without COVID-19 diagnosis received both tests for screening for SARS-CoV-2, of whom 12 were tested positive for SARS-CoV-2 by RT-PCR tests in their throat swabs and were removed from the analyses. In total, we included 1470 patients with COVID-19, 3832 healthcare providers, 19555 workers, and 1616 other patients in the final analyses (N=26473). Follow up time for death among hospitalized COVID-19 patients was calculated as from symptom onset until either discharge from hospital or April 15, 2020, which came first.

Diagnosis of COVID-19 was based on epidemiological history, clinical manifestations and presence of SARS-CoV-2 in clinical samples confirmed by using real-time RT-PCR method.^11^ There were changes in diagnosis of COVID-19 in China, and the case definition was gradually broadened to allow for detection of milder cases.^49^ The confirmed cases were estimated to be 4 times less than that if the later broader case definition had been adopted earlier. Severity of status of patients with COVID-19 at admission was defined as moderate, severe, or critical. Patients with mild diseases were not admitted to the above three hospitals and were generally admitted to Fangcang Hospitals (makeshift hospitals).

### RT-PCR test for SARS-CoV-2 virus RNA

Clinical specimens collection and RT-PCR test for SARS-CoV-2 were previously described^18^. Clinical specimens in COVID-19 patients included nasal swabs, throat swabs, sputum, anal swabs, and bronchoalveolar lavage (BAL), and clinical specimens in healthcare providers without confirmed COVID-19 diagnosis were only throat swabs. In brief, clinical specimens were collected from these people by trained nurses or physicians wearing proper personal protection equipment. RT-PCR tests for SARS-CoV-2 were performed using a nucleic acid detection kit following the manufacturer’s protocol. The test simultaneously amplifies and detects two target genes, including open reading frame 1ab (ORF1ab) and nucleocapsid protein (N). Primers used for those two target genes are as follows: ORF1ab: forward primer CCCTGTGGGTTTTACACTTAA, reverse primer ACGATTGTGCATCAGCTGA; and the probe 5′-VIC-CCGTCTGCGGTATGTGGAAAGGTTATGG-BHQ1-3′; N: forward primer GGGGAACTTCTCCTGCTAGAAT, reverse primer CAGACATTTTGCTCTCAAGCTG, and the probe 5′-FAM-TTGCTGCTGCTTGACAGATT-TAMRA-3′. Conditions for the amplifications were incubation at 50 °C for 15 minutes and 95 °C for 5 minutes, followed by 40 cycles of denaturation at 94 °C for 15 seconds and extension at 55 °C for 45 seconds. The diagnostic criteria for positive and negative RT-PCR results were based on the recommendation by the National Institute for Viral Disease Control and Prevention (China): positive result <37 cycle threshold value (Ct-value) and negative result ≥40. A Ct-value of 37-39 required retesting.

### COVID-19 IgM/IgG test for antibodies against SARS-CoV-2

Serum samples from these people were collected. Methods for testing serum IgM and IgG antibodies to SARS-CoV-2 were previously described.^50^ COVID-19 IgM/IgG test kits contained recombinant SARS-CoV-2 antigens (spike protein and nucleocapsid protein) labelled with magnetic beads (tested on a fully ⍰ automated chemiluminescence immunoassay analyzer) or colloidal gold (test card), anti-human IgM monoclonal antibody, and anti-human IgG monoclonal antibody. These test kits were reported to have high sensitivity and specificity^27,50^. According to the manufacturers, the sensitivity and specificity are ∼90% and >99% for IgM, and ∼98% and ∼98% for IgG, respectively.

Two physicians extracted the following data using data collection form from electronic medical records: demographic information such as age and sex, RT-PCR test date and results, COVID-19 IgM/IgG test date and results, date of symptom onset for COVID-19 patients, treatments received, and clinical outcomes. Another physician in the research team reviewed the collected data.

### Statistical Analysis

Continuous variables were reported using mean and 95% confidence interval (CI) if normally distributed or median and interquartile if nonnormally distributed. Categorical variables were described as frequency rates and percentages. The χ2 test was used for the comparison of categorical variables and Fisher’s exact test was used when frequency was too low. Multigroup comparisons were performed using ANOVA test, following by Tukey test for adjusting for multiple comparisons. Prevalence of positive IgG test results and 95% CI was also reported. For the assessment of RT-PCR test results of SARS-CoV-2 and IgM/IgG test results, the last test result for each person was used in the analyses. Kaplan-Meier curves were plotted to show survival differences in hospitalized COVID-19 patients by the status of IgG antibodies to SARS-CoV-2. Adjusted hazard ratio and 95% CI were calculated by fitting a Cox proportional hazard model, controlling for age, sex, and severity of COVID-19 at admission. Statistical analyses were conducted using SAS software version 9.4 (SAS Institute; Carey, NC). A 2-sided p value of <0.05 was considered statistically significant.

## Data Availability

Data and analyses codes are available from the corresponding authors on request. All request for raw and analyzed data and materials will be reviewed by the corresponding authors to verify whether the request is subject to any intellectual property or confidentiality obligations. Access will be granted after a signed data access agreement is attained.

